# COVID-19 pandemic: Analyzing of spreading behavior, the impact of restrictions and prevention measures in Germany and Japan

**DOI:** 10.1101/2021.04.22.21255953

**Authors:** Stefan Bracke, Alicia Puls, Masato Inoue

## Abstract

In December 2019, the world was confronted with the outbreak of the respiratory disease COVID-19. The COVID-19 epidemic evolved at the beginning of 2020 into a pandemic, which continues to this day. The incredible speed of the spread and the consequences of the infection had a worldwide impact on societies and health systems. Governments enforced many measures to control the COVID-19 pandemic: Restrictions (e.g. lockdown), medical care (e.g. intensive care) and medical prevention (e.g. hygiene concept). This leads to a different spreading behavior of the COVID-19 pandemic, depending on measures. Furthermore, the spreading behavior is influenced by culture and geographical impacts. The spreading behavior of COVID-19 related to short time intervals can be described by Weibull distribution models, common in reliability engineering, in a sound way. The interpretation of the model parameters allows the assessment of the COVID-19 spreading characteristics. This paper shows results of a research study of the COVID-19 spreading behavior depending on different pandemic time phases within Germany and Japan. Both countries are industrial nations, but have many differences with respect to historical development, culture and geographical conditions. Consequently, the chosen government measures have different impacts on the control of the COVID-19 pandemic. The research study contains the analyses of different pandemic time intervals in Germany and Japan: The breakout phase in spring 2020 and subsequently following waves until winter season 2020/2021.

## 1. Introduction

In December 2019, the world was confronted with the outbreak of the respiratory disease COVID-19. The COVID-19 epidemic evolved at the beginning of 2020 into a pandemic, which continues to this day. The incredible speed of the spread and the consequences of the infection had a worldwide impact on societies, health systems and social life. Governments enforced many measures to control the COVID-19 pandemic: Restrictions (e.g. distance regulations, shutdown of educational system and breakdown of public traffic system), medical care (e.g. further development of intensive care) and medical prevention (e.g. provisioning of masks, hygiene concept). This leads to a different spreading behavior of the COVID-19 pandemic, depending on country or region, cf. Bracke et al. (2020). Furthermore, the spreading behavior is influenced by culture and geographical impacts. The spreading behavior of COVID-19 related to short time intervals can be described by Weibull distribution models, common in reliability and safety engineering, in a sound way. The interpretation of the model parameters (shape and location parameter; cf. Sec. 3) allows the assessment of the COVID-19 spreading characteristics.

This paper shows results of a research study of the COVID-19 spreading behavior depending on different pandemic time phases within Germany and Japan.

On the one side, both countries are industrial nations and have experience to deal with pandemics (e.g. 1918-1919 Spanish flu, 1957–1958 Asian flu pandemic, 1968 flu pandemic (Hong Kong flu), 2009-2010 pandemic H1N1 (swine flu)). Some of the genes were equal respectively similar, therefore, the Spanish flu is called “mother of all pandemics”. Germany and Japan have both made constant efforts to facilitate water and sewage plant, improve the housing environment, expand immunization, and develop antibacterial drugs to rid of infectious diseases. As a result, the number of people dying from infectious diseases, e.g. such as tuberculosis and pneumonia, decreased sharply in the middle of the 20th century.

On the other side, Germany and Japan have many differences with respect to historical development, culture and geographical conditions. Consequently, the chosen restrictions, medical care and prevention measures have different effectiveness respectively impacts on the control of the COVID-19 pandemic. The research study contains the analyze of different pandemic time intervals in Germany and Japan: The breakout phase in spring 2020, first wave and lockdown, second and third wave in autumn 2020 respectively winter season 2020/2021. Furthermore, aspects from the differences in German and Japanese culture, social life and geographic are considered.

## 2. Goal of Research Study

The overarching goal is analyses of the COVID-19 spreading behavior depending on different pandemic time phases within Germany and Japan; cf. Sec 5. The detailed analyses are as follows:

1. Comparison of COVID-19 spreading behavior characteristics within first, second and third wave.
2. Analyze of the impact of the chosen measures (restrictions and prevention) on the control of the COVID-19 pandemic.
3. Discussion of differences of both countries with respect to historical development, culture and geographical conditions

The analyses were performed using reliability engineering methods and technical statistics methods, cf. Sec. 3. Base of operations are data and information from Johns Hopkins University data Base, cf. Sec. 4.

## 3. Method

This section shows the statistic fundamentals for analyzing and comparing the COVID-19 pandemic in Germany and Japan. The spreading behavior in the different pandemic waves and the impact of measures like lockdown is analyzed by using the Weibull distribution model, cf. Sec. 3.1. The detection of the different waves is made with a Cox-Stuart trend test (Significance test), cf. Sec. 3.2.

### 3.1. Weibull distribution model

The two-parameter Weibull distribution model is given based on Eq. (1), cf. Weibull (1951).

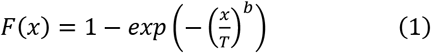

The parameters, besides the term life span variable x, are scale parameter T (in lifetime analysis: characteristic life span) and shape parameter b. By variating parameter b, different failure rates can be described, therefore the Weibull model can be flexibly used for different applications, cf. Rinne (2008). The shape parameter b gives hints regarding the character of the failure period: early failure period, random failure or operation time related failure behavior. The scale parameter T indicates the x value of the probability 0.633%. The Weibull parameters are estimated by using the Maximum Likelihood Estimator (MLE), cf. Fisher (1912).

The applicability of this distribution model for the analyses of occurrence of infection is given by the exponential increase of number of cases of COVID-19. The Weibull distribution model offers the possibility to gain knowledge with regard to the infection development in comparison to classical methods of virology like SIR model (cf. Kermack and McKendrik (1927)) or the basic reproduction number. The easy interpretability of the Weibull parameters allows the analysis of the spreading behavior, in particular the spreading speed. The central thinking transfer is the interpretation of the shape parameter b, the gradient of the Weibull distribution model (log-log-scale), as spreading speed. This is the first advantage in comparison to the use of an exponential distribution model. Second advantage is the normalization of the Weibull distribution function: It allows an easy comparison of measurement data based on different time ranges (samples). Therefore, the Weibull distribution model with the corresponding parameters and probability plots is the base for the comparison of the different COVID-19 waves in Germany and Japan. (Puls and Bracke 2020)

### 3.2. Cox & Stuart trend test

The Cox Stuart trend test is a non-parametric statistical test for detecting trends in a sample, based on the Binomial distribution. The data is divided in the midpoint into two sequences and the paired difference D is build. For the detection of the second wave, the one-sided form of the test is used to determine an upward trend. Therefore, the number of the positive signs in D is defined as S+. The null hypothesis states that S+ follows a binomial distribution with the number of experiments n as number of elements of D and a probability 0.5. If the p-value of the test is smaller than the significance level alpha, the null hypothesis is rejected and an uptrend is confirmed; cf. Cox and Stuart (1955):

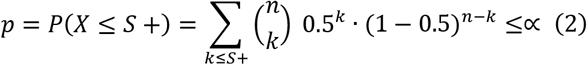

For the detection of the different COVID-19 waves the daily confirmed cases are analyzed as a time series. A one-sided trend test (upward trend) is performed with 14 data points and a significance level α of 0.05, cf. Eq. 2. The tested hypotheses are as follows, cf. Puls and Bracke (2020):

- Null hypothesis: There is no upward trend.
- Alternative hypothesis: There is an upward trend (second wave).

The sample size of 14 days is chosen to mitigate outliners and data falsifications, cf. Sec. 4.2. This trend test is repeated until the whole period under review is analyzed. Besides these test decisions, the development of number of cases is taken into account for the differentiation of the pandemic waves.

## 4. Data base and uncertainty

In this section, the data base for the analyses and comparison of the different COVID-19 waves is described (Sec. 4.1). Also, uncertainty factors are mentioned and the handling with these data falsification is outlined, cf. Sec. 4.2.

### 4.1. Data base

In Germany, the COVID-19 data is collected by the Robert Koch Institute (RKI). The laboratory confirmed cases are documented in combination with several pandemic key figures, cf. RKI (2021). The national data source for the COVID-19 cases in Japan is the Ministry of Health, Labor and Welfare, cf. MHLW (2021).

To enable a comparison of Germany and Japan in the different COVID-19 pandemic phases, a unified data base is necessary. Therefore, instead of using the national data sources, the data documentation of the Johns Hopkins University (JHU) is used. The COVID-19 dashboard of this university documents confirmed cases, recovered cases as well as death cases regarding countries and regions, starting at 01-22-2020, cf. JHU (2021). Hence, an international data base with the same data collection methods for all countries is given.

### 4.2. Data uncertainty

It must be considered, that the data quality within the JHU database is different, caused by reasons with relation to the reported countries: E.g., data can be incomplete and censored, depending on the collection and reporting system of the country. Furthermore, the different definitions of facts (e.g. death case: death with COVID-19 or because of COVID-19) has an impact on the data. This section gives a brief overview of the uncertainty with regard to data acquisition, for detailed explanations cf. Bracke et al. (2020).

First of all, there are the criteria for testing. Differences in the test strategy e.g. symptom based or area-wide affect the confirmed number of cases. In Japan, there are two cases of testing: symptom-based testing and testing without reason. In the first case, the doctor determined that the subject needs a PCR test due to the symptoms. In the second case, applicants can undergo PCR tests at private laboratories if they bear the costs even if they have no symptoms. In Germany, in the first pandemic phase, there was only the symptom-based testing due to the determination by a doctor. In a later pandemic phase, there is the possibility for all citizens to do one pay-free rapid test in test centers per week. When this test gets positive, a PCR test is determined. The positive rapid tests do not enter the data base of COVID-19, only the PCR confirmed cases define the number of cases in Germany.

Secondly, different reporting systems have to be taken in account. In Japan, the reporting procedure goes from the National Institute of Infectious Diseases, over the quarantine station to the local institutions for health to the public health institutes until the data of the number of cases accessed the official reporting center by the Ministry of Health, Labor and Welfare. In Germany the numbers of suspected diseases, diseases and death in relation to COVID-19 are reported. The report is made to the responsible health department by doctors, members of other medical or nursing professions and heads of institutions like nursing homes within 24 hours.

Distortions in the data basis due to accessibility of health department also must be taken into account as an uncertainty factor with respect to the interpretation of the spreading behavior analysis. As well in Japan as in Germany the public health institutes are closed at weekend, so there is a weekend impact in the data base. During the weekends, the reported number of cases decreases and after the weekend there are more cases due to the delay of reporting.

In general, apart for the political measures and differences in measurement, there are many other factors that influence the virus spread and thus the data base. Some of these uncertainty factors are (without claiming to be conclusive), cf. Dimmock et al. (2016):

- Seasonality and climatic effects, cf. Sajadi et al. (2020)
- Frequency of susceptible individuals in the population, like urbanity and persons in agglomerations (population density)
- Differences in behavior, e.g. cultural or climatic determined
- Type of treatment, cf. Gattinoni et al. (2020)

The concrete impact of these points for the comparison of the different pandemic phases in Japan and Germany are analyzed and discussed in detail in Sec. 5.2.2.

With the knowledge that these uncertainty factors occur, the comparative data is kept as constant as possible. Therefore, besides the use of the unified data base of JHU, the following aspects are concerned:

- The data is ranked and the time is normalized by the date of the first infection in the particular pandemic phase.
- The data is differentiated between the pandemic phases under and without measure impact.
- For the comparison of different phases and for the comparison of the spreading in Germany and Japan the same time span for all phases and countries is used.

Furthermore, all results are checked for plausibility and uncertainties during the analyses.

## 5. COVID-19 spreading behavior: Data Analytics

This section focuses on COVID-19 data analytics. At first, the different pandemic phases with measure impact are analyzed country by country, cf. Sec. 5.1.1 resp. 5.1.2. The different waves are detected with Cox-Stuart Trend test, the spreading behavior is analyzed by the Weibull distribution models and the different measures are evaluated. In the second step, the spreading of COVID-19 in the different waves is compared for Germany and Japan, cf. Sec. 5.2. An overview of the number of cases and the corresponding Weibull distributions are given in Sec. 5.2.1, while in Sec. 5.2.2 the reasons for the differences are discussed.

### 5.1. Spreading behavior per country

At first, the development of COVID-19 in Germany is analyzed, afterwards the spreading behavior of COVID-19 in the different waves in Japan is estimated.

#### 5.1.1. Overview of COVID-19 in Germany

Before the different pandemic phases can be analyzed with Weibull distribution models, a differentiation of the waves of COVID-19 has to be made. Therefore, the Cox-Stuart Trend test is conducted for the daily confirmed cases in Germany, as described in Sec. 3.2. As a result, the p values (cf. Eq. (2)) are plotted in Fig. 1 on the right ordinate. For comparison, the corresponding daily confirmed cases are deducted on the left ordinate. The corresponding dates are assigned to these values. Additionally, the significance (α = 0.05) is shown as a horizontal red line. The period under review for the two waves of COVID-19 in Germany are marked with blue resp. green vertical lines.

**Fig. 1.**
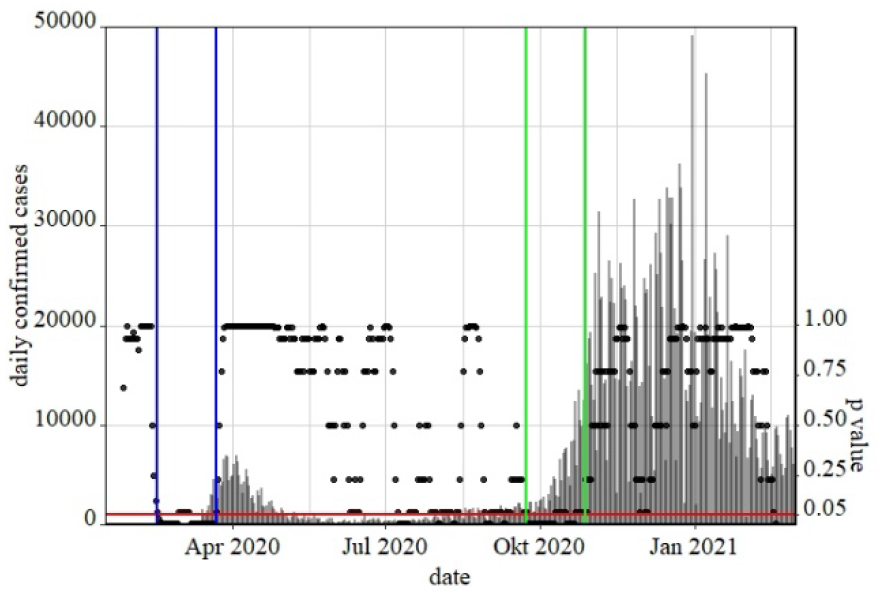
Detection of waves of COVID-19 in Germany with Cox-Stuart trend test, p values (black points), α=0.05 (red line), daily confirmed cases (grey lines), first wave (blue lines) and second wave (green line).

All points under the red line represent those tests, which result in an upward trend. When the p value is below the significance level, the null hypothesis is rejected and the alternative hypothesis of an upward trend is assumed, cf. Sec. 3.2. According to this approach, the start of the first wave in Germany was on 02-16-2020, marked with the first vertical blue line in Fig. 1. The first lockdown in Germany was on 03-22-2020. As the first wave should represent the unhindered spreading as base for the comparison and the evaluation of the measure impact, the end of the first wave is set to 03-21-2020. This results in a data base of 35 days, five weeks. For a reasonable comparison, this time span should be the period under review for all pandemic phases analyzed in the further.

Analogous the second wave is detected by the analyze of the results of the Cox-Stuart trend test and the development of the daily confirmed cases. The corresponding time span in marked by two vertical green lines in Fig. 1. Overall, there are five different pandemic phases in Germany: first wave, first lockdown, second wave, lockdown light and second lockdown. The different characteristics of these phases especially of the lockdown measures are described in the following, here an overview of the relevant time spans for the Weibull distribution models is given, cf. Table 1.

**Table 1.** COVID-19 development in Germany. Important phases and dates of the periods under review for the Weibull analyses.

| phase | start date | end date |
| --- | --- | --- |
| first wave | 02-16-2020 | 03-21-2020 |
| first lockdown | 03-22-2020 | 04-25-2020 |
| second wave | 09-23-2020 | 10-27-2020 |
| lockdown light | 11-02-2020 | 12-06-2020 |
| second lockdown | 12-16-2020 | 01-29-2021 |

Note: The start dates of the first and second wave are detected with Cox-Stuart trend test. The start date of the lockdown measures are the dates of entry into force of these measures. The end dates are defined by the given time span of 35 days of each pandemic phase in the analyses.

In Germany, the first COVID-19 case was reported on 01-28-2020. Middle February 2020 the number of cases exponentially increased. First measures like prohibition of major events or shutdown of educational systems and retail or border controls were taken middle March 2020, before there was the first lockdown measure on 03-22-2020. Additional to the first measures, there were distance regulations in this phase.

After the pandemic has subsided in summer 2020, the second wave came in September 2020. The German government reacted with a “lockdown light”, a measure with prohibition of mayor events, distance and contact restrictions and mask obligation in November 2020. This measure was expanded to the second lockdown middle December 2020 with a shutdown of the educational system and retail as well as border controls.

To compare the spreading behavior in these different pandemic phases, Weibull distribution models are fitted to the corresponding ranked data. Results are the Weibull probability plot (log-log scale) in Fig. 2 and the Weibull parameters with confidence belts documented in Table 2.

**Table 2.** Weibull model parameters COVID-19 in Germany. Different pandemic phases with lockdown (LD) measures (cumulative confirmed cases). Confidence level γ = 0.95.

| phase | Cases | scale T [d] | shape b, confidence belt |
| --- | --- | --- | --- |
| first wave | 22,215 | 32 - | $11.80 \leq 11.92 \leq 12.05$ |
| first LD | 134,300 | 17 | $1.669 \leq 1.677 \leq 1.684$ |
| second wave | 186,007 | 27 | $2.949 \leq 2.960 \leq 2.971$ |
| LD light | 650,204 | 20 | $1.673 \leq 1.676 \leq 1.680$ |
| second LD | 680,529 | 18 | $1.520 \leq 1.523 \leq 1.526$ |

**Fig. 2.**
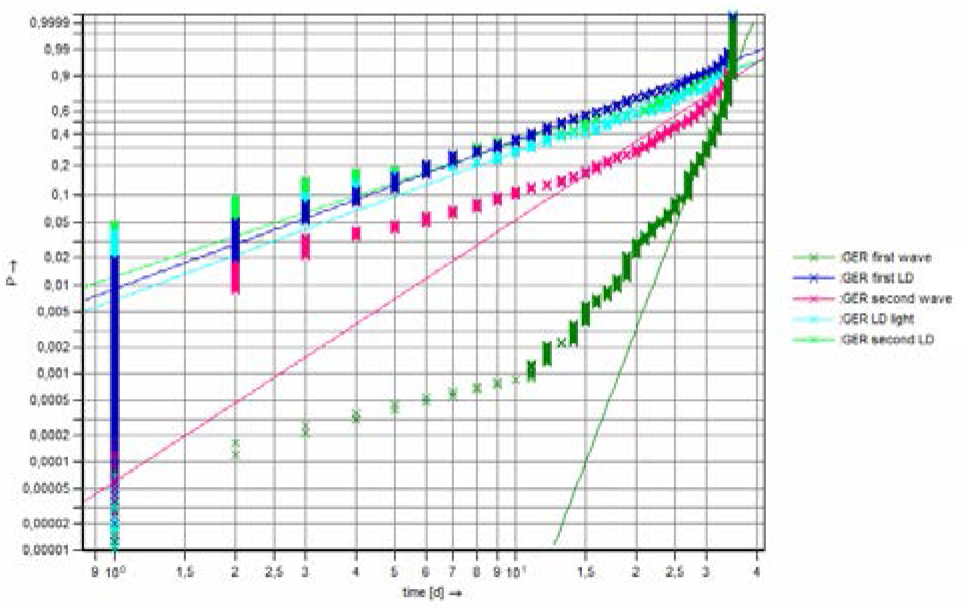
Weibull distribution models COVID-19 in Germany (GER), comparison of different pandemic phases with lockdown (LD) measures, confirmed cases, time span 35 days.

With the Weibull distribution models, an analysis of the spreading behavior is possible. By comparing the slope of the curves in Fig. 2 is gets clear, that the spreading speed in the first and second wave without lockdown measures is much higher than with lockdown measures. The shape parameter (resp. spreading speed) in the first pandemic phase in Germany is factor 7 higher than the spreading speed under the first lockdown. It stands out, that the spreading speed in the first wave is higher than in the second wave. This is explainable with the measures in force in the second wave (e.g. masks, distance regulations), which were executed in the first wave.

The spreading speed during the different lockdown measures is on a similar level. The lowest spreading speed was in the second lockdown, the shape parameters in first lockdown and lockdown light are not significant different. Noticeable is the difference in the scale parameter T between the first resp. second wave and the spreading during the lockdown measures. With lockdown measures, the scale parameter of the Weibull distribution models is significantly lower. With a probabilitiy of P = 0.633, the infections were earlier in the period under review, consequently with P = 0.367 the infections are distributed on a longer time span than in the first and second wave. This is another indicator, that the lockdown measures effectively decreased the spreading speed of COVID-19 in Germany.

#### 5.1.2. Overview of COVID-19 in Japan

Analogical to the approach of the analysis of the spreading behavior of COVID-19 in Germany, for the analysis of Japan at first the Cox-Stuart trend test is done to detect the different pandemic phases, cf. Fig. 3. Again, the p values are plotted in combination with the daily confirmed cases. The significance α = 0.05 is represented by a red line, the first (blue), second (green) and third (yellow) waves are delimited with vertical lines. Deliberately in each case the beginning of the phase is chosen in order to get the data base of the exponential increase for the Weibull analyses.

**Fig. 3.**
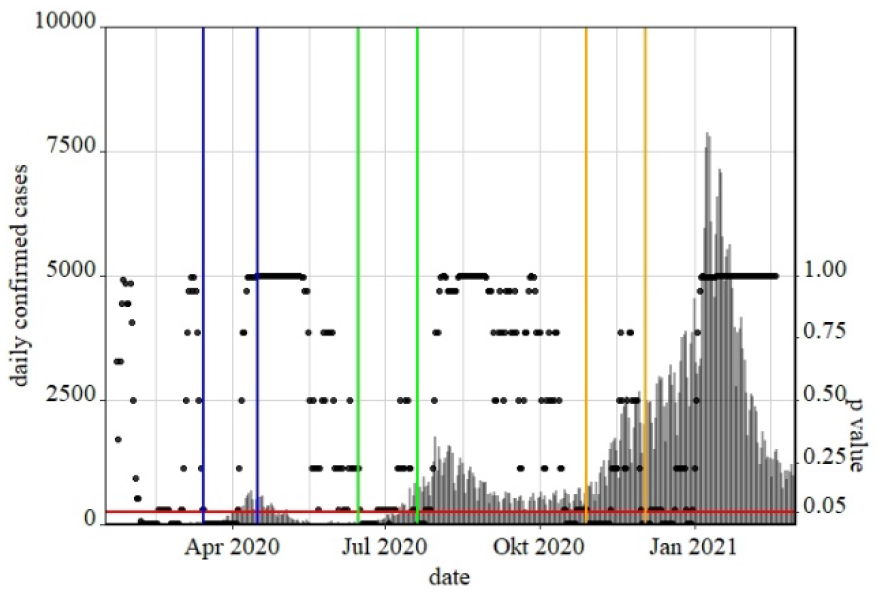
Detection of waves of COVID-19 in Japan with Cox-Stuart trend test, p values (black points), α=0.05 (red line), daily confirmed cases (grey lines), first wave (blue lines), second wave (green line) and third wave (yellow line).

In every wave there was one specific measure. Hence, there are six different pandemic time spans for the Weibull analyses, documented in Table 3.

**Table 3.** COVID-19 development in Japan. Important phases and dates of the periods under review for the Weibull analyses.

| phase | start date | end date |
| --- | --- | --- |
| first wave | 03-15-2020 | 04-15-2020 |
| first state of emergency | 04-16-2020 | 05-20-2020 |
| second wave | 06-15-2020 | 07-19-2020 |
| “go to travel” campaign | 07-22-2020 | 08-25-2020 |
| third wave | 10-29-2020 | 12-02-2021 |
| second state of emergency | 01-08-2021 | 02-11-2021 |

Note: The start dates of the first, second and third wave are detected with Cox-Stuart trend test. The start date of the measures are the dates of entry into force. The end date of the first wave is determined by the begin of the first state of emergency. The other end dates are defined by the given time span of 35 days of each pandemic phase in the analyses.

In Japan, the first confirmed COVID-19 infection was on 01-16-2020. As a first reaction, the elementary school, junior high school and the high school were closed on 02-27-2020. Further measures were the declaration of a state of emergency for big cities followed by the request to warning area regarding home office and to refrain from eating and drinking establishments at night. On 04-16-2020 there was a declaration of a state of emergency for the whole country, which is the period under review for the central measure in the first pandemic phase in Japan.

The second wave began middle June 2020 and took until September 2020. The main measure in this pandemic phase was the “go to travel” campaign, which should revive the tourism industry in combination with measures to decrease the virus spreading. There were distance regulations, mask obligation and hygiene measures.

The third pandemic wave started in the end of October 2020 and reached its maximum in middle of January 2021. As the governmental reaction the second state of emergency was declared for whole of country on 01-08-2021 with several measures:

- Request to avoid unnecessary outings, especially stay at home request after 8 pm
- Limitation of events (up to 5,000 people and less than 50% of the capacity until 8 pm)
- Request to refrain from eating and drinking establishments until 8 pm
- Promotion of telework (home office); 70% reduction in the number of employees

In contrast to the first declaration of emergency, there was no temporary school closing.

To compare the spreading behavior in the different pandemic phases, Weibull distribution models are fitted to the corresponding ranked data. Results are the Weibull probability plot (log-log scale) in Fig. 4 and the Weibull parameters with confidence belts documented in Table 4.

**Table 4.** Weibull model parameters COVID-19 in Japan. Different pandemic phases with different measures (cumulative confirmed cases). Confidence level γ = 0.95

| phase | cases | scale<br>T [d] | shape b,<br>confidence belt |
| --- | --- | --- | --- |
| first wave | 8,040 | 26 - | $4.117 \leq 4.190 \leq 4.264$ |
| first state of<br>emergency | 7,486 | 12 | $1.292 \leq 1.316 \leq 1.340$ |
| second<br>wave | 8,077 | 28 | $3.426 \leq 3.487 \leq 3.549$ |
| “go to<br>travel”<br>campaign | 37,425 | 20 | $1.920 \leq 1.936 \leq 1.952$ |
| third wave | 54,555 | 24 | $2.427 \leq 2.444 \leq 2.460$ |
| second state<br>of<br>emergency | 145,041 | 15 | $1.451 \leq 1.457 \leq 1.463$ |

**Fig. 4.**
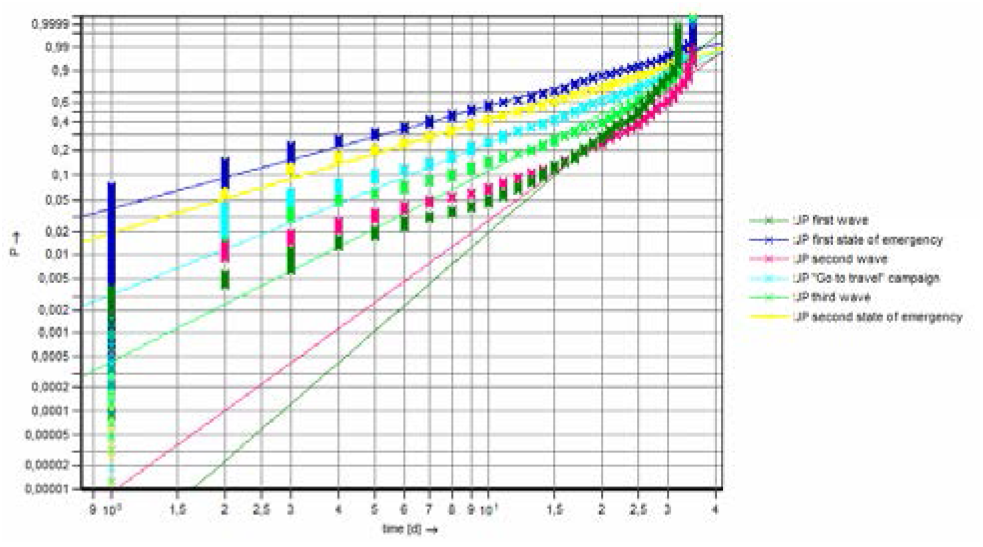
Weibull distribution models COVID-19 in Japan (JP), comparison of different pandemic phases with different measures, confirmed cases, time span 35 days.

Analyzing the slope of the curves (representing the spreading speed) in Fig. 4, differences between the phases of first, second and third wave in comparison to the different measure phases get clear. With the governmental measures the spreading speed is lower than the spreading speed of the waves with (nearly) unhindered development. This difference is significant due to the comparison of the shape parameters with confidence belts in Table 4. For example, the first state of emergency reduced the spreading speed of the first wave with factor ∼3.2, while the second state of emergency decreased the spreading speed of the third wave with factor ∼1.7. Notifiable is also the fact, that the scale parameter T is lower in the pandemic phases with concrete measures. Analogous to the analyses of the spreading behavior in Germany this underlines the efficiency of these measure to reduce the spreading speed.

In addition, a downward trend in the spreading speed from first to second to third wave is in evidence. During the different pandemic phases, the spreading speed slows down. One reason could be the awareness in the population or the measures in force. Regarding the different phases with specific measures the following ranking is identifiable regarding the efficiency of slowing down the pandemic spreading: the first state of emergency followed by the second state of emergency followed by the “go to travel” campaign. The daily number of cases in contrast increased during the first year of the pandemic in Japan, especially during the third wave the daily confirmed cases reached a peak.

### 5.2 Comparison of the spreading behavior in the different pandemic waves in Germany and Japan

In this section, at first the spreading behavior of COVID-19 in Germany and Japan is compared with data analytics. The numbers of cases are opposed and the Weibull distribution models are analyzed, cf. Sec. 5.2.1. In a second step, the results are discussed based on differences in culture, geography and pandemic strategies, cf. Sec. 5.2.2.

#### 5.2.1 Overview of comparison COVID-19 in Germany and in Japan

The first difference which gets clear are the different number of cases of COVID-19 in Germany and Japan. In Fig. 5 the daily confirmed cases are plotted over the time for both Germany (red) and Japan (blue).

**Fig. 5.**
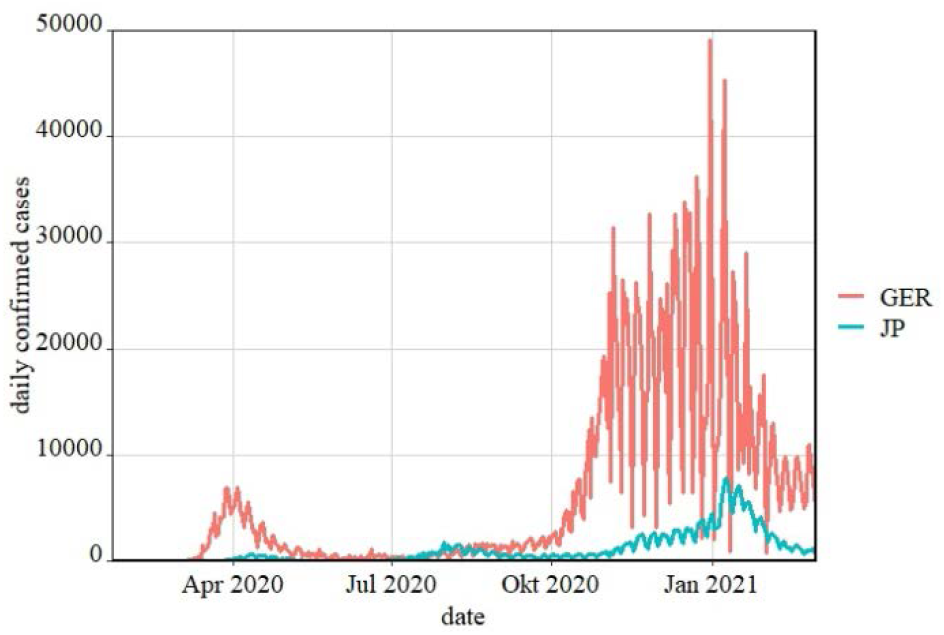
Daily confirmed cases in Germany (GER) and Japan (JP) January 2020 until March 2021.

Both in the first third of 2020 and in the third third of 2020, in Germany there were much more cases (factor ∼5) than in Japan though Japan has more inhabitants (∼126 Mio.) than Germany (∼83 Mio.). Additionally, the occurrence of infection differs. The first waves were in both countries approximately at the same time. Japan had a second wave in summer 2020, when the spreading was subsided in Germany. In middle September the second wave started in Germany, while in Japan the third wave came end October 2020. These differences represent the different development of the virus in Europe and Asia.

For a detailed comparison of the spreading behavior of COVID-19 in Germany and Japan, the Weibull distribution models of the different waves are merged in one plot, cf. Fig. 6. The corresponding Weibull parameters are documented in Table 5.

**Table 5.** Weibull model parameters COVID-19 in Germany (GER) and Japan (JP). Different pandemic waves (cumulative confirmed cases). Confidence level γ = 0.95.

| phase | cases | scale<br>T [d] | shape b,<br>confidence belt |
| --- | --- | --- | --- |
| first wave<br>GER | 22,215 | 32 - | $11.80 \leq 11.92 \leq 12.05$ |
| second<br>wave GER | 186,007 | 27 | $2.949 \leq 2.960 \leq 2.971$ |
| first wave<br>JP | 8,040 | 26 - | $4.117 \leq 4.190 \leq 4.264$ |
| second<br>wave JP | 8,077 | 28 | $3.426 \leq 3.487 \leq 3.549$ |
| third wave<br>JP | 54,555 | 24 | $2.427 \leq 2.444 \leq 2.460$ |

**Fig. 6.**
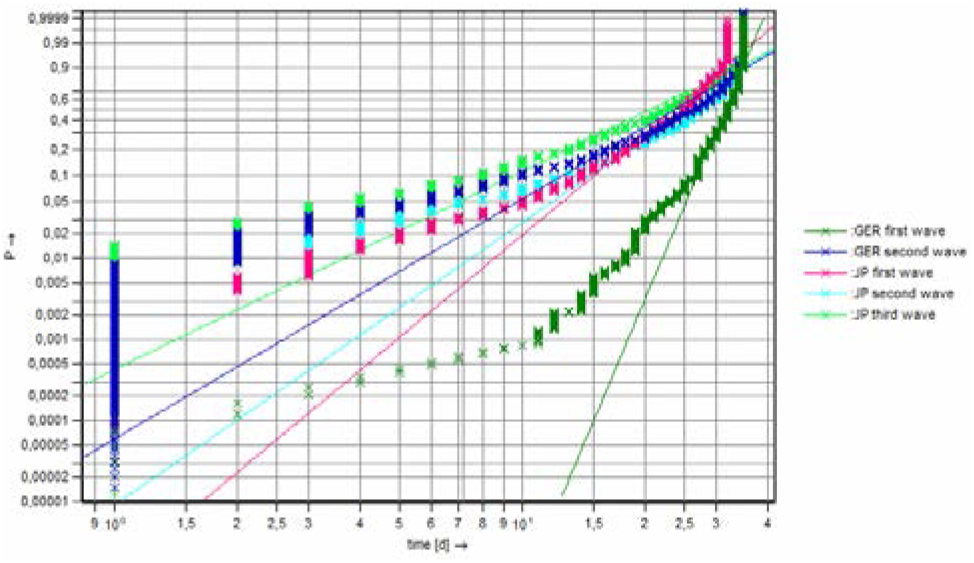
Weibull distribution models COVID-19 in Germany (GER) and Japan (JP), comparison of different pandemic waves, confirmed cases, time span 35 days.

Regarding the first wave, the spreading speed (resp. slope, resp. shape parameter) in Germany was ∼3 times greater than in Japan. The second wave in Germany is on a comparable level with the third wave in Japan regarding the spreading speed. During the second wave in Japan, the spreading speed was greater in comparison to Germany.

The impacts of the different measures in Germany and Japan are opposed with the Weibull distribution models in Fig. 7 and Table 6.

**Table 6.** Weibull model parameters COVID-19 in Germany (GER) and Japan (JP). Different pandemic measures/lockdown (LD) (cumulative confirmed cases). Confidence level γ = 0.95.

| measure | cases | scale<br>T [d] | shape b,<br>confidence belt |
| --- | --- | --- | --- |
| first LD GER | 134,300 | 17 | $1.669 \leq 1.677 \leq 1.684$ |
| LD light GER | 650,204 | 20 | $1.673 \leq 1.676 \leq 1.680$ |
| second LD<br>GER | 680,529 | 18 | $1.520 \leq 1.523 \leq 1.526$ |
| first state of<br>emergency JP | 7,486 | 12 | $1.292 \leq 1.316 \leq 1.340$ |
| “go to travel”<br>campaign JP | 37,425 | 20 | $1.920 \leq 1.936 \leq 1.952$ |
| second state<br>of emergency<br>JP | 145,041 | 15 | $1.451 \leq 1.457 \leq 1.463$ |

**Fig. 7.**
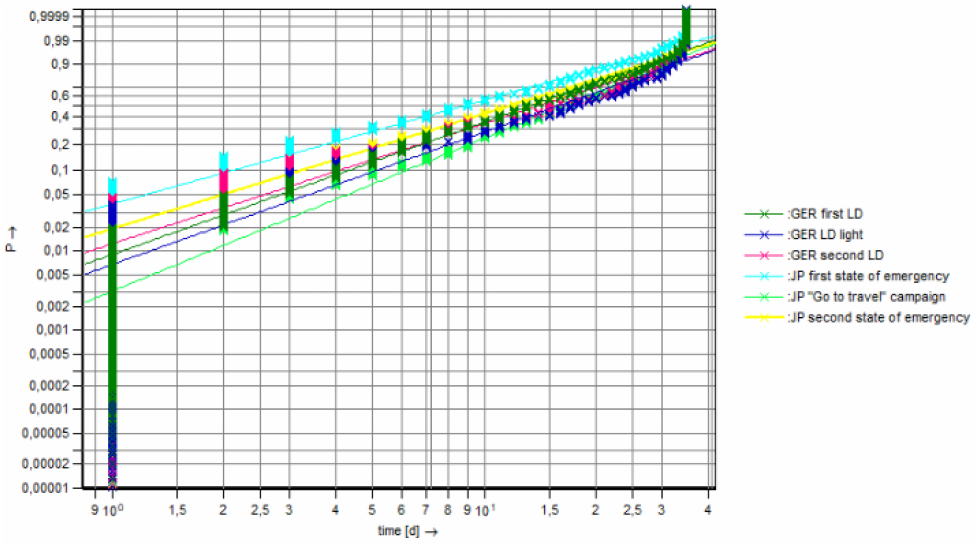
Weibull distribution models COVID-19 in Germany (GER) and Japan (JP), comparison of different pandemic measures/lockdown (LD), confirmed cases, time span 35 days.

The spreading speed of all pandemic phases with measures is at a comparable level as can be seen on the similar slope of the curves in Fig. 7. The states of emergency in Japan were significant more effective to reduce the spreading speed than the lockdown measures in Germany. The greatest shape parameter was during the “go to travel” campaign in Japan. It is notifiable, that the measures in Germany decreased the occurrence of infection percentual more effective than the measures in Japan, cf. Table 5 in comparison to Table 6. Altogether the number of cases and the spreading speed of COVID-19 is lower in Japan than in Germany.

#### 5.2.2 Discussion

As explained in Sec. 4.2 several uncertainty factors can affect the analyses of the spreading behavior of COVID-19. Concluding the data analytics, here the main differences in these aspects between Germany and Japan are described as a base of operations for the interpretation of the results in the previous section. The following Table 7 shows the main aspects of uncertainty with the particular characteristic for Germany resp. Japan without claiming to be conclusive.

**Table 7.**
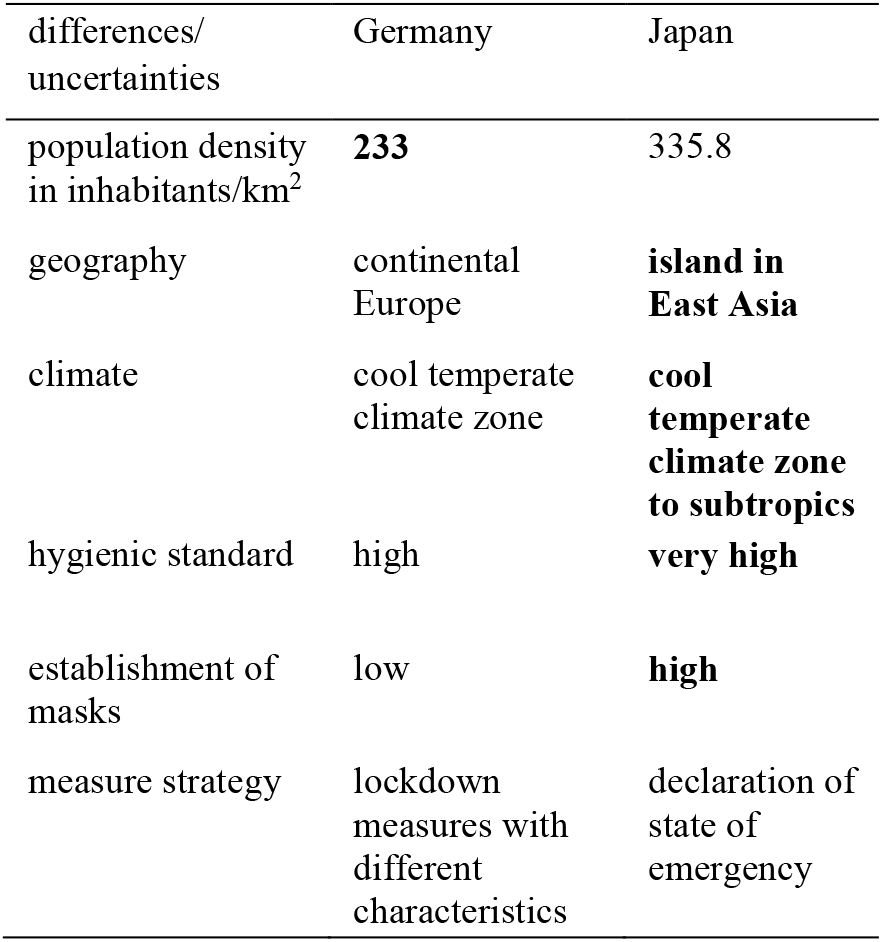
Differences/ uncertainties in the comparison of the spreading behavior of COVID-19 in Germany and Japan. Highlighted the better basis conditions for a slow virus spreading speed.

| differences/<br>uncertainties | Germany | Japan |
| --- | --- | --- |
| population density<br>in inhabitants/km<sup>2</sup> | **233** | 335.8 |
| geography | continental<br>Europe | **island in**<br>**East Asia** |
| climate | cool temperate<br>climate zone | **cool**<br>**temperate**<br>**climate zone**<br>**to subtropics** |
| hygienic standard | high | **very high** |
| establishment of<br>masks | low | **high** |
| measure strategy | lockdown<br>measures with<br>different<br>characteristics | declaration of<br>state of<br>emergency |

With exception from the population density Japan has advantageous basis conditions for a slow virus spreading speed. The geographical position as island enables a strict entry check. Additionally, the culturally based very high hygienic standard in combination with the establishment of wearing masks in the public (before the COVID-19 pandemic) stunt the virus spreading. These factors in combination explain the slow spreading speed of Japan in the first wave in comparison to the spreading speed of Germany in the first wave and the low number of cases of Japan in the first half of 2020.

Regarding the measure strategy due to the similar level of spreading speed (cf. Table 6) neither the lockdown strategy of Germany nor the state of emergency strategy of Japan can be proved as more efficient. While the first German lockdown measure reduced the spreading speed percentual the most, the first state of emergency in Japan lead to the lowest spreading speed resp. shape parameter, cf. Table 6.

## 6. Summary

In December 2019, the world was confronted with the outbreak of the respiratory disease COVID-19. The incredible speed of the COVID-19 spreading behavior and the consequences of the infection had a worldwide impact on societies, health systems and social life. This paper shows results of a research study of the COVID-19 spreading behavior depending on different pandemic time phases within Germany and Japan. The impact of the enforcement of measures to control the COVID-19 pandemic, like restrictions (e.g. lockdown and medical prevention (e.g. hygiene concept) are compared within the different COVID-19 waves in Germany and Japan.

As an overall result can be stated: Within the time phases of states of emergency in Japan were significant more effective to reduce the spreading speed in comparison to the time phase of the lockdown measures in Germany. Comparing the different waves of COVID-19 in Germany and Japan it gets clear that for both countries the greatest spreading speed was during the first wave. In first wave, the spreading speed was about factor 3 higher in Germany than in Japan. Reasons can be the high hygienic standard or the establishment of masks before the pandemic in Japan. The second and third waves in both countries are on a similar level regarding the spreading behavior. Notifiable is the difference in the number of cases: Germany has much greater number of cases of COVID-19, one reason can be the different geographic position (continental Europe vs. island).

Concerning the different measures, it can be stated, that all measures significantly reduced the spreading speed. The percentual greatest reduction (∼ factor 7) was during the first lockdown in Germany. The lowest spreading speed of all analyzed measures was during the first state of emergency in Japan. The other measures were on a comparable level in both countries regarding the spreading behavior. Measures like shutdown of educational system or retail, wearing masks of telework effectively slow down the occurrence of infection.

## Data Availability

All data are from COVID-19 dashboard of Johns Hopkins University (JHU)

https://gisanddata.maps.arcgis.com/apps/opsdashboard/index.html#/bda7594740fd40299423467b48e9ecf6

